# Characterizing Potential Interaction Between Respiratory Syncytial Virus and Seasonal Influenza in the U.S.

**DOI:** 10.1101/2023.10.04.23296424

**Authors:** Jiani Chen, Deven V. Gokhale, Liang Liu, Pejman Rohani, Justin Bahl

## Abstract

RSV and seasonal influenza are two of the most important causes of respiratory infection that consistently peak during winter months in the U.S. Here, we characterized the circulation of these viruses in the U.S. with weekly positive case reports and genetic surveillance and used a mathematical modeling approach to explore their potential interaction at an HHS regional level. Our analyses showed RSV and seasonal influenza co-circulate with various relatively epidemic sizes and seasonal overlaps across seasons and regions. We found RSV might have different evolutionary dynamics compared to seasonal influenza, with local persistence may play a role in underlying annual epidemics. Our analysis supports a competitive interaction between RSV and seasonal influenza in most HHS regions and we speculate that cross-immunity after infection might be the major driver of viral competition. Together, our work supports the competition between RSV and seasonal influenza across the U.S. at a population level. Our findings are important for the future development of protective strategies against these respiratory viruses.

## Introduction

Interaction among infectious diseases has been very well documented in various polymicrobial disease systems [1]. In the context of respiratory infections, a well-known example is the association between the influenza virus and the bacterium pneumococcus, driven by enhanced susceptibility to secondary bacterial colonization subsequent to influenza infection [2][3][4]. Another important example comes from the 2009 influenza A pandemic (H1N1pdm09), with data from several European countries indicating that the spread of the virus might have been interrupted by the annual autumn rhinovirus epidemic [5]. These pathogen-pathogen interactions may lead to cooperation or competition, and their co-occurrence within the same host population can profoundly alter the pathogenesis and spread of infectious diseases with substantial public health consequences [6].

Respiratory Syncytial Virus (RSV) and seasonal influenza virus (Flu) are two of the most important causes of respiratory infection [7][8][9]. Both RSV and seasonal influenza activity consistently peak during winter months in the US. However, the relative disease burden and similarity between the epidemic timing of these viruses vary substantially among regions and seasons [9], [10]. The burden of influenza varies from season to season, depending in part on the dominant virus type or subtypes in circulation [11]. Influenza subtypes A/H3N2 and A/H1N1 (Flu A) have become the leading cause of seasonal influenza illness and death in the U.S. over the last 50 years [12]. Additionally, two distinct lineages of the influenza B (Flu B) virus, Victoria and Yamagata, co-circulate or alternate with Flu A and have received greater attention in recent years [13]. Intensive studies of the molecular evolution of the influenza virus have provided important insights into its seasonal emergence and spread in human populations [11][14][12][13]. Similar to seasonal influenza, two distinct antigenic subgroups of RSV have been identified, RSV-A and RSV-B, which show clear phylogenetic divergence. Genotype ON (RSV-A) and BA (RSV-B) are the two most prevalent RSV genotypes that are circulating in the U.S. in recent years [15][16], [17]. However, the molecular epidemiology of RSV is largely unknown both on local and global scales [18]. Since RSV and seasonal influenza have similar seasonality and clinical presentation, researchers often try to extrapolate knowledge gleaned from the study of seasonal influenza to RSV to understand viral dynamics and develop protective strategies against these respiratory viruses-induced illnesses together [19]. Therefore, elucidating the exact nature of the interaction between RSV and seasonal influenza can greatly enhance our ability to forecast future epidemics, and substantially inform the development and implementation of control strategies.

Both biological and epidemiological studies suggest potential competitive interaction between RSV and seasonal influenza [20]. The proposed biological mechanism for the competition between these viruses is the activation of the innate “antiviral response” by an infection that can inhibit further or subsequent infection of another virus, resulting in a period of cross-protection during or after infection [18], [21]. Epidemiological data show that when rates of infection with RSV are high, influenza infections are low; the converse is also true [22]. The interaction between respiratory viruses has the potential to impact virus genetic diversity and epidemics pattern and therefore would lead to the observed patterns in phylogenetics or surveillance data [11], [25]. Previous studies have demonstrated the application of dynamic modeling to test biological hypotheses on pathogen interaction mechanisms and it could be used to infer the interaction between RSV and seasonal influenza [23]. However, very few studies examine the potential interaction between RSV and seasonal influenza on a population level, and the explicit mechanism of their interaction has not been well-studied.

In this study, we initially carry out a phylogenetic analysis to elucidate differences in the evolutionary trajectories of RSV and seasonal influenza. We then apply a seasonally forced, two-pathogen, mechanistic transmission model to explicitly investigate the nature of ecological interference between RSV and seasonal influenza and quantify its effects in driving transmission dynamics in different regions in the U.S. Our analysis provides evidence of the potential negative interaction of RSV and seasonal influenza on the ecological scale and the cross-immunity after infection might be an important mechanism.

## Results

### Seasonality overlaps between RSV and seasonal influenza

Different relative epidemic sizes and peaking times of RSV and seasonal influenza can be observed across different HHS regions in the U.S. (SI Appendix, Fig. S1, represented regions are shown in Fig.1A). We computed the correlation coefficient of these viruses using weekly surveillance reports and found the positive case of RSV and seasonal influenza increased with another (Fig.1B, SI Appendix, Table S1), suggesting epidemic seasonality similarities between these viruses in the U.S. In addition, the magnitude of the correlation between RSV and seasonal influenza are various across HHS regions, with a relatively high value in HHS Regions 6, 7, and 10 between RSV and Flu A and in HHS regions 6, 7, and 8 between RSV and Flu B. We further computed the time-lagged correlation of different pairs of viruses (Fig.1C) to measure the temporal association between RSV and seasonal influenza epidemics in the U.S. In our analysis, we observed the maximize correlation with zero to little (positive/negative) time lags in different regions, indicating the seasonality overlap between RSV and seasonal influenza are different across regions and might have different interaction patterns.

**Fig. 1.**
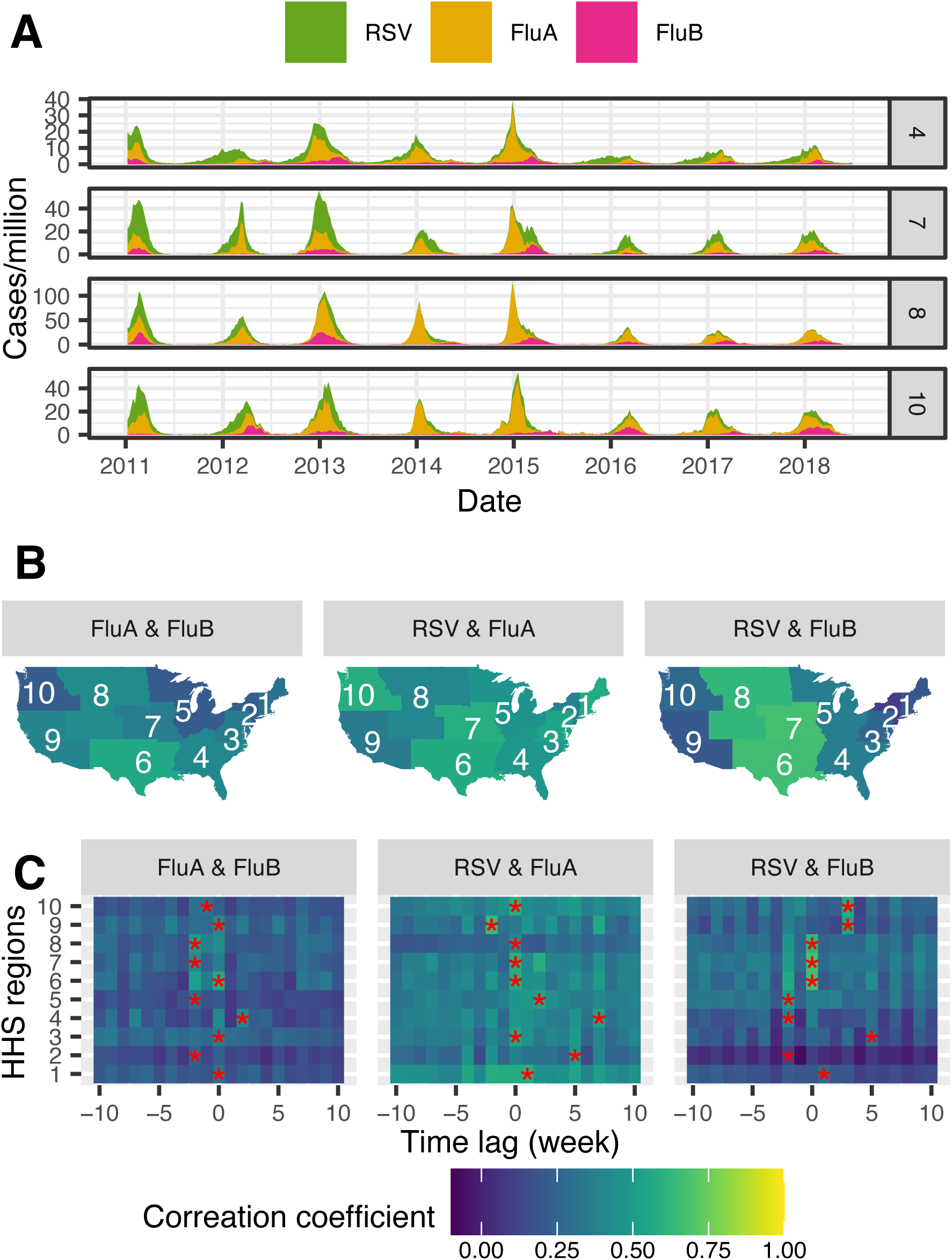
Correlation coefficient of RSV and seasonal influenza epidemics in the U.S. **A.** Weekly RSV and seasonal influenza epidemics in represented HHS regions (HHS Region 4,7,8,10). **B.** Correlation coefficient between RSV and seasonal influenza weekly surveillance reports (Table S1). Relative color in HHS regions indicates the normalized correlation coefficient of weekly positive samples of the different pairs of viruses. **C**. Time-lagged correlation coefficient between RSV and seasonal influenza (Table S2). The time lags (week) that maximize the correlation coefficient in each HHS region are labeled with *. Negative values indicate a leading correlation and positive values indicate a lagging correlation. Lighter-colored indicate a higher correlation coefficient between two viruses and darker-colored indicate a lower correlation coefficient.

### Population dynamics of RSV and seasonal influenza

We characterized the population dynamics of RSV and seasonal influenza using a phylogenetic approach. The phylogenies of seasonal influenza lineages were estimated using Hemagglutinin (HA) gene segments that were collected in the U.S. during the 2011-2019 seasons. The analyses of RSV were based on G gene sequences from northern hemisphere countries in the same period and we focus on genotypes ON and BA. We calculated the time-sliced statistics (Materials and Methods, TMRCA: Time to the most recent ancestor, and Diversity: branch distance between pairs of tips) from RSV and seasonal influenza phylogenies (Fig. 2A). A periodic reduction in TMRCA and diversity across time can be observed from seasonal influenza phylogenies (H1, H3, Victoria, and Yamagata). In contrast, TMRCA and diversity scores of RSV phylogenies increased across time, except for the analysis of BA in recent seasons, which might be due to a lack of RSV sampling during recent sampling times. In addition, Bayesian “Skyride” coalescent reconstructions demonstrate strong periodicity of seasonal influenza viruses, which are correlated with their seasonal epidemic patterns, though we observed very low genetic diversity of Flu B lineages in some years. The population dynamics of RSV did not show seasonal fluctuation and both ON and BA genotypes presented peaks from 2011 to 2019 (Fig.2B, Fig.2C). These different phylogenetic patterns suggest local persistence may play a role in underlying annual epidemics of RSV and influenza strains are under constant negative selection.

**Fig. 2.**
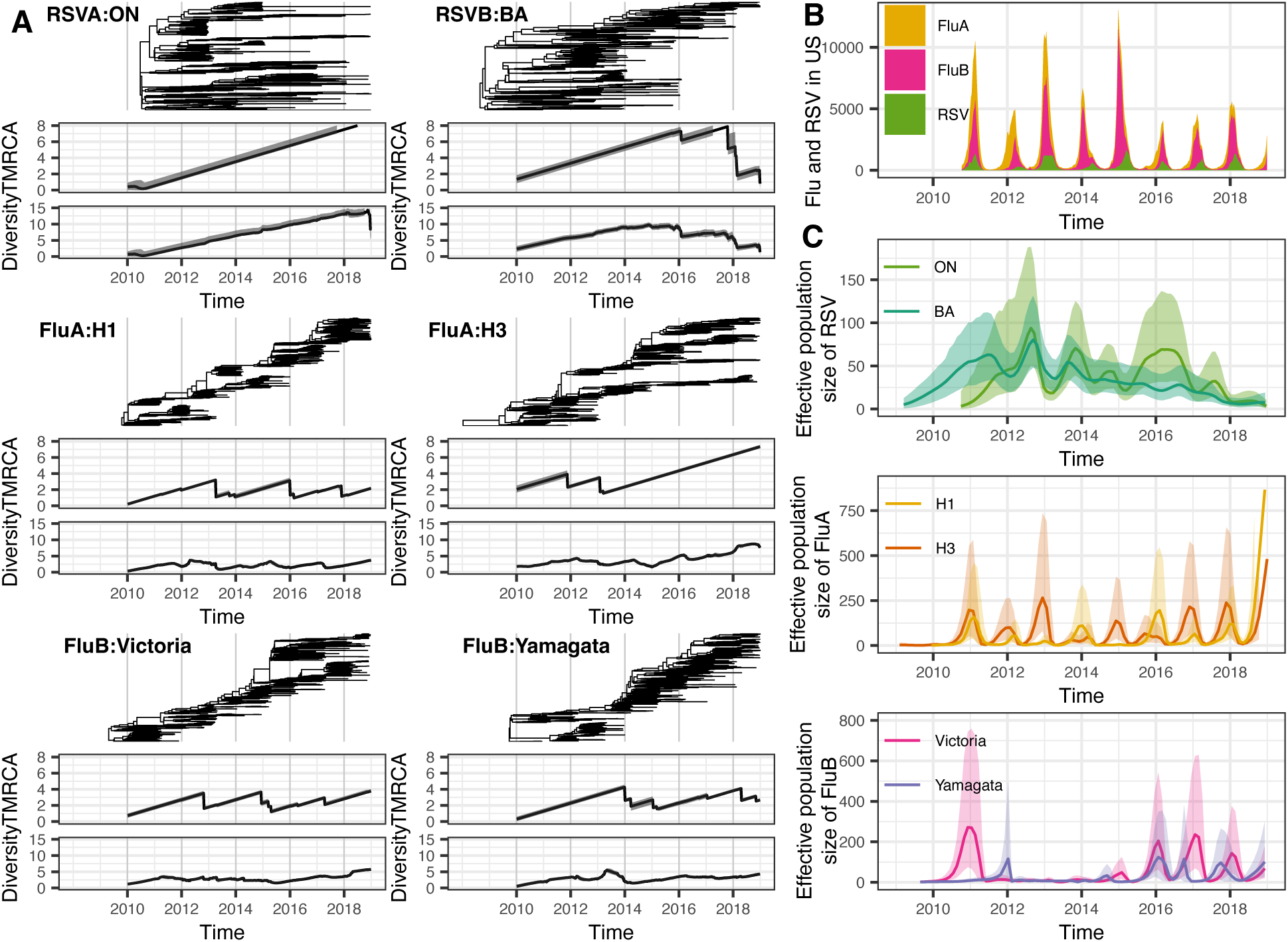
Population dynamics of RSV and seasonal influenza in the U.S. **A.** Phylogenies and corresponding time-slice statistics. The time-slice TMRCA and Diversity are calculated by breaking the phylogeny into multiple temporal sections. TMRCA, time to the most common ancestor of all tips within temporal sections. Diversity is the average time to coalescent for pairs of lineages within temporal sections. TMRCA and Diversity are measured in the unit of years. Solid lines represent mean values and gray outlines represent 95% CI across MCMC replicates. **B.** Surveillance curve and reconstructed effective population size of RSV and seasonal influenza using GMRF Bayesian “Skyride” analysis. Solid lines represent mean values and outlines represent 95% CI across MCMC replicates. We computed the correlation coefficient using these estimated time series effective population sizes of RSV and seasonal influenza to explore their potential association at the virus population level (SI Appendix, Table S2). Significant negative correlation coefficients can be observed between RSV and seasonal influenza strains: RSV-A genotype ON with influenza H3 (CI: −0.513, −0.128), H1 (CI: −0.482, −0.087), and Victoria (CI: – 0.482, −0.087); RSV-B genotype BA with influenza H1 (CI: −0.506, −0.138) and Yamagata (CI: −0.564, −0.217), which indicate these viruses might have a negative association.

### Potential competition between RSV and seasonal influenza

The co-circulation of RSV and seasonal influenza suggests a transmission model that allows the co-infection of these viruses is required. Therefore, we adapted a two-pathogen transmission model to investigate the potential competition between RSV and seasonal influenza (SI Appendix Fig.S4, Table S3, Table S4) in the U.S. This model accommodates two mechanisms of competition between pathogens. Competition may arise through the inhibition of co-infection, which is modeled by scaling the transmission rates of secondary infections with parameter 𝜓. In addition, following infection with one pathogen, individuals may gain short-term cross-immunity against another pathogen and can be similarly modeled by scaling the force of secondary infection by 𝜒. Changing the value of these two parameters can influence the timing, magnitude, and shape of observed epidemics (Fig. 3).

**Fig. 3.**
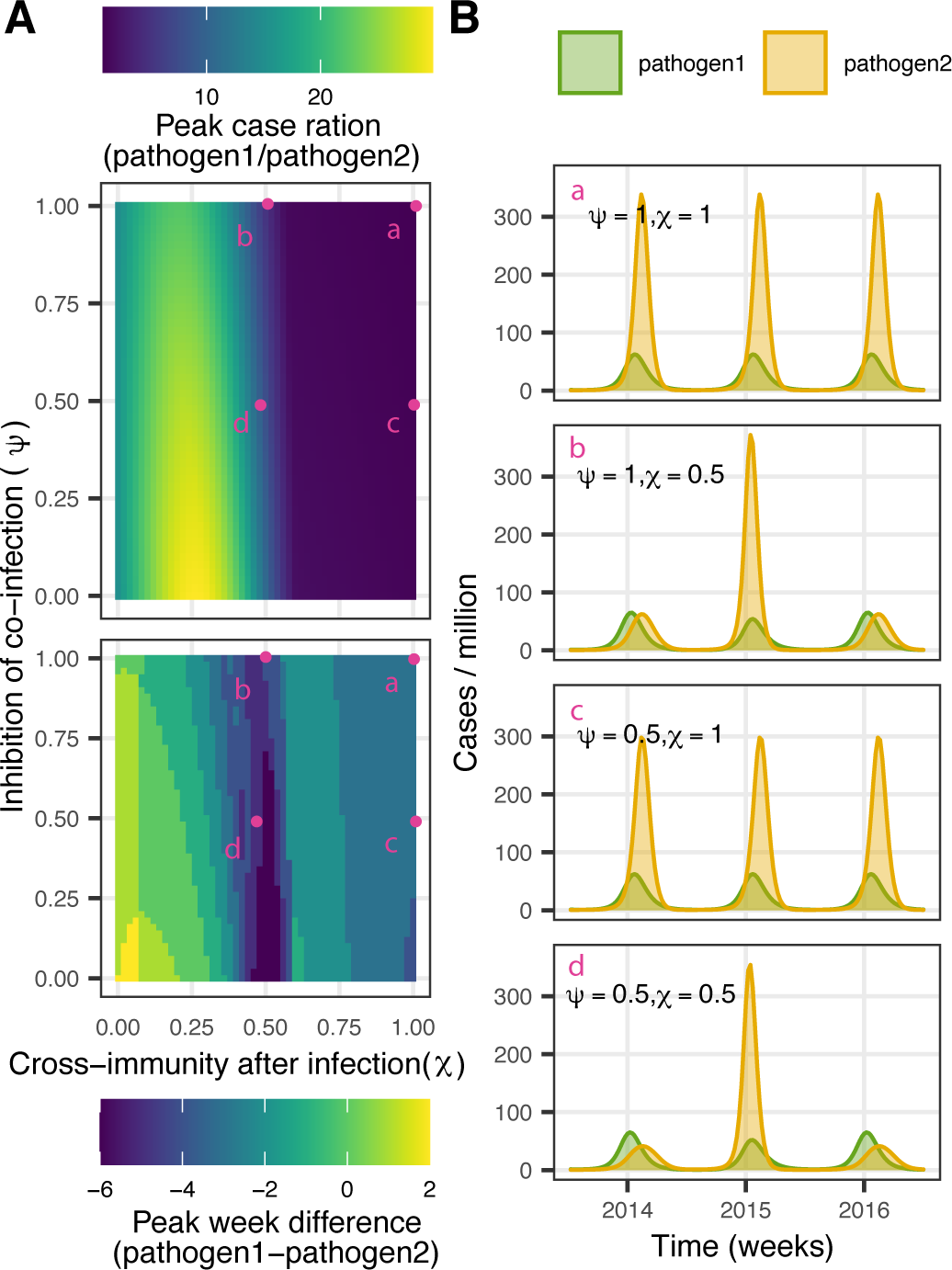
Simulation study to examine the effects of the proportion of inhibition of co-infection (𝝍) and cross-immunity after infection (𝝌) using a two-pathogen transmission model. **A.** Changes in relative peak case ratio and the relative time of peaks based on the values 𝜓 and 𝜒 in the model. **B.** Simulated trajectories resulting from parameter values of 𝜓 and 𝜒 selected at points a, b, c, and d in A.

We fit this model to RSV and type-specific seasonal influenza (Flu A or Flu B) incidence data that are collected from 10 HHS Regions from 2014 to 2017 and use Akaike Information Criterion (AIC) values to compare four different hypotheses regarding the interaction between RSV and seasonal influenza: (1) no interaction (𝜒 = 𝜓 = 1); (2) inhibition of co-infection (𝜓 < 1, 𝜒 = 1); (3) transient cross-immunity after infection (𝜓 = 1, 𝜒 < 1); and (4) both the inhibition of co-infection and transient cross-immunity (𝜓 < 1, 𝜒 < 1)(SI Appendix Table S5). In these analyses, the duration of short-lived cross-protection is fixed at 6 months [24]. We estimated parameters that could optimize the likelihood estimates of the model under each hypothesis in 10 HHS regions (SI Appendix Table S7 to Table S16). The virus-specific simulated trajectories under four hypotheses for 10 HHS regions are shown in SI Appendix, Fig. S5. We found that adding competitive interaction between RSV and seasonal influenza provides a better explanation of the surveillance data in most regions except for HHS Region 8 (SI Appendix, Table S6), and the epidemic trajectories from surveillance data in different HHS regions can be well fit under the corresponding best hypothesis as examined by Root Mean Square Error (RMSE) (SI Appendix, Fig. S6). As described in Table 1, different levels of cross-immunity after infection were estimated across HHS regions under the corresponding best hypothesis, and the inhibition of co-infections was also supported in some regions (HHS Regions 1, 3, 4, 5, 6, 7 for RSV and Flu A, HHS Regions 3, 5, 6, 10 for RSV and Flu B). We used HHS Region 1 as an example region to demonstrate our findings: our analysis suggests RSV and Flu A are fully inhibited to be co-infected and there was also a short-term cross-immunity, with a 45% reduction in the infection rate of another pathogen. By fitting the RSV and Flu B surveillance, we found a 39% reduction in the infection of the second virus, and the inhibition of co-infection was not supported (Fig. 4, Table 2).

**Fig. 4.**
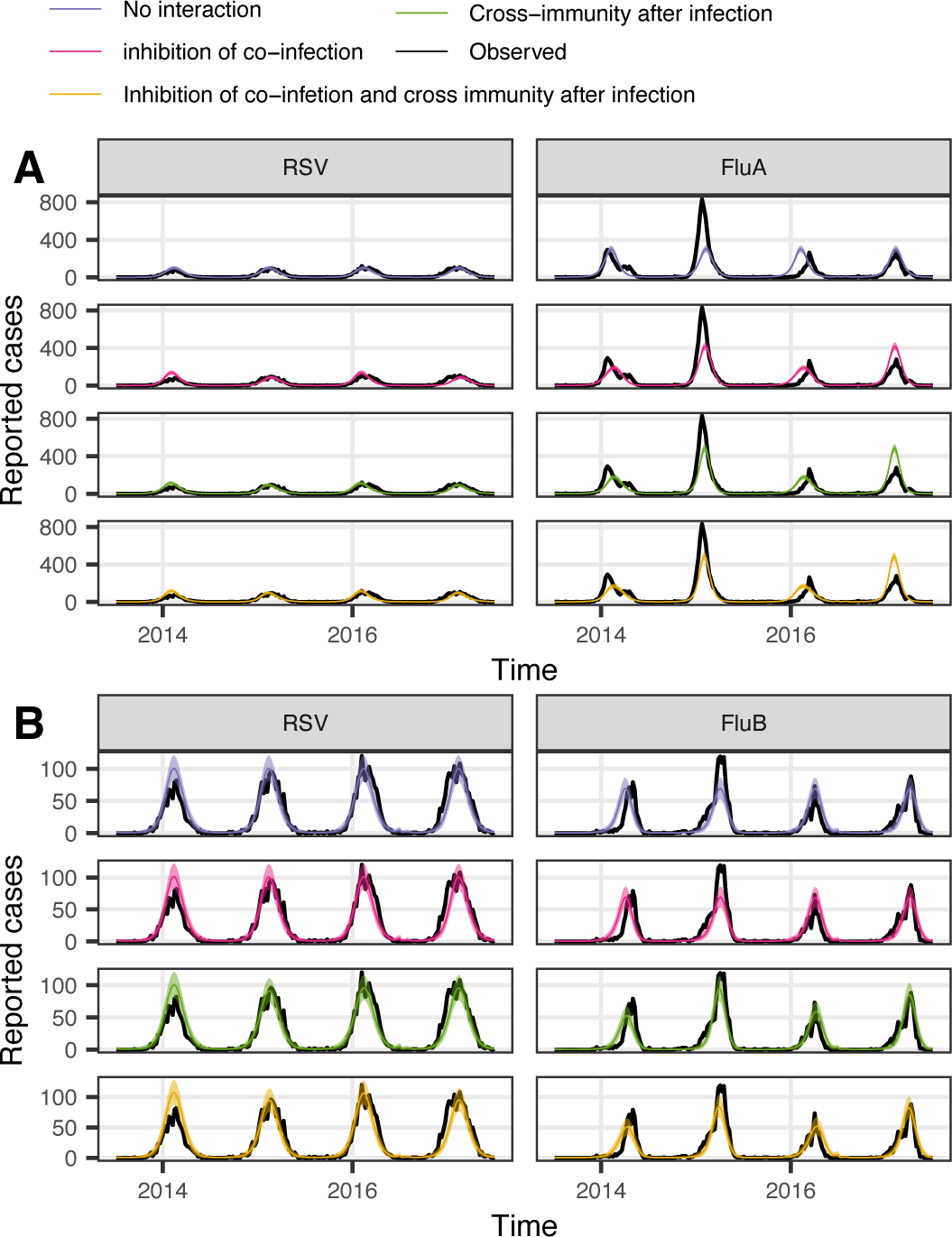
Relative fits of four epidemiological hypotheses for season 2014-2017 in HHS Region 1. Matched virus-specific simulated trajectories under four hypotheses are shown in different colors (Mean in solid line and 95% CI in shading). The black solid line represented the HHS regional level weekly case report data. **A.** The test of the interaction between RSV and Flu A. **B.** The test of the interaction between RSV and Flu B.

**Table 1.** Competition parameter estimates under the best fit hypothesis for seasons 2014-2018 in 10 HHS regions.

| HHS region | RSV-Flu A |  | RSV-Flu B |  |
| --- | --- | --- | --- | --- |
| | Inhibition of co-infection($\psi$) | Cross-immunity after infection($\chi$) | Inhibition of co-infection($\psi$) | Cross-immunity after infection($\chi$) |
| 1 | 0 | 0.55 | 1 | 0.64 |
| 2 | 1 | 0.69 | 1 | 0.72 |
| 3 | 0 | 0.59 | 0 | 0.8 |
| 4 | 0.57 | 0.37 | 0 | 0.87 |
| 5 | 0 | 0.78 | 1 | 0.61 |
| 6 | 0 | 0.77 | 1 | 0.45 |
| 7 | 0 | 0.52 | 0.95 | 0.64 |
| 8 | 1 | 1 | 1 | 1 |
| 9 | 1 | 0.79 | 0.2 | 0.6 |
| 10 | 1 | 0.5 | 1 | 0.2 |

**Table 2.**
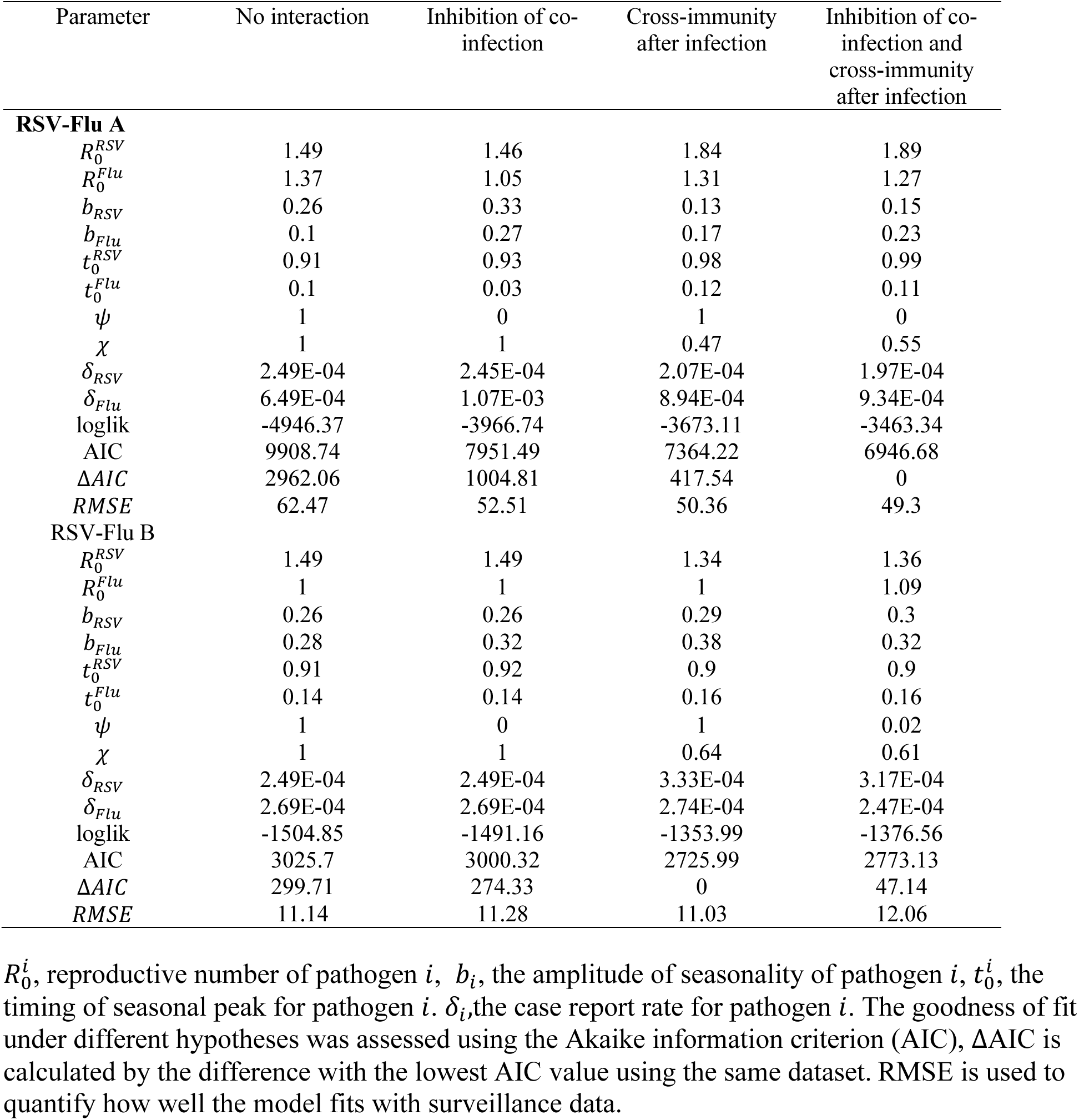
Parameters estimates and epidemiological hypotheses evaluation for season 2014-2017 in HHS regions 1.

We further estimated the credible interval (CI) of the two competitive parameters under the best-fit hypothesis in 10 HHS regions (SI Appendix, Fig. S7). As shown in the analysis of HHS Region 1 (Fig. 5), changing the values of 𝜓 have relatively small effects on the likelihood profile compared with the strength of cross-protection (𝜒), and the CI of 𝜒 can be well identified (RSV and Flu A: 0.522, 0. 567, RSV and Flu B: 0.628, 0.638). Our results indicate that short-term cross-immunity after the infection might be the major mechanism to explain the competition between RSV and seasonal influenza.

**Fig. 5.**
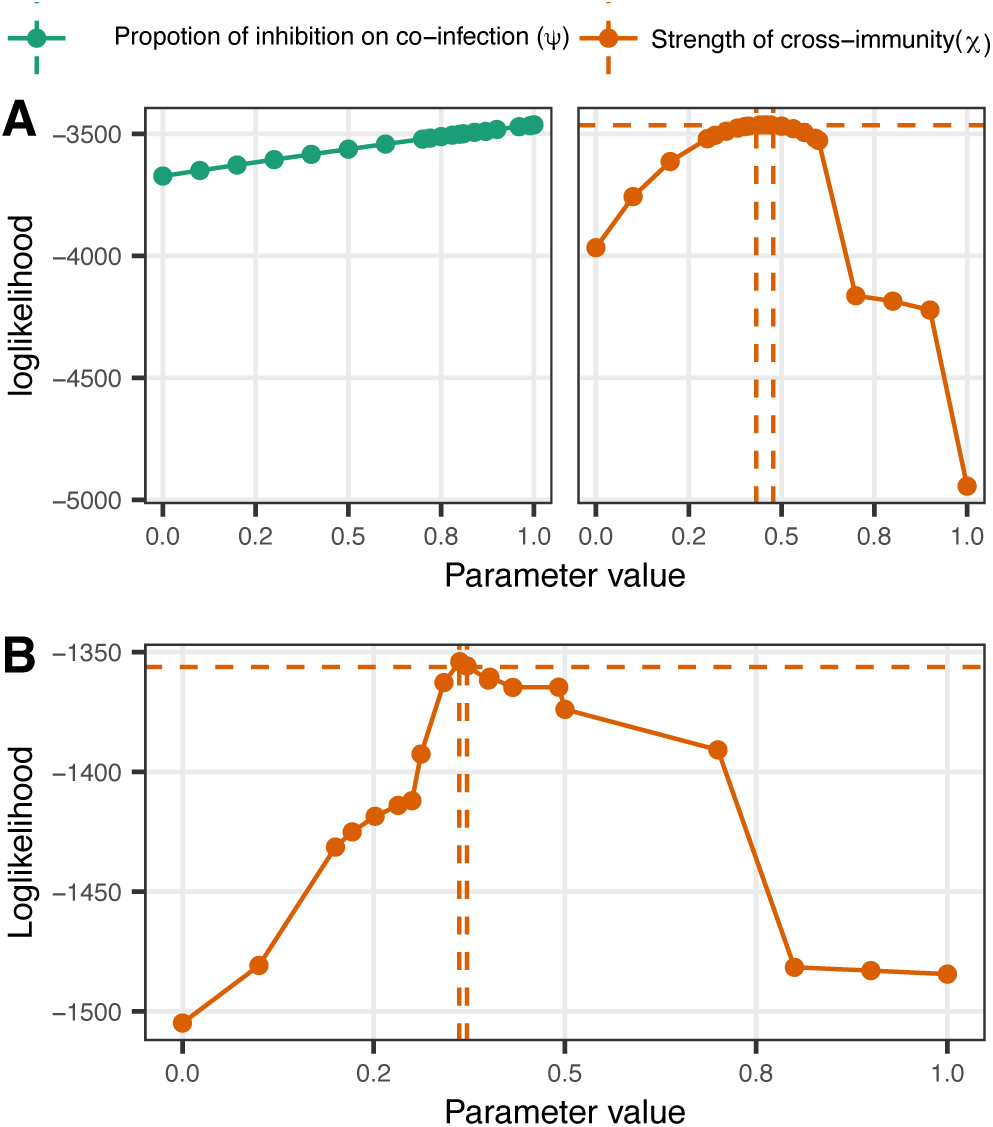
Likelihood profile tests of competition interaction parameters for RSV and seasonal influenza, inferred in HHS Region 1 from 2014 to 2017. Plotted in each graph is the likelihood profile for the inhibition of co-inhibition (𝜓) in green and the cross-immunity after infection (𝜒) in orange, which are lines connected by repeated likelihood estimates (N =20, shown in colored solid circles). The values within the two dashed lines are within the estimated 95% CI. **A.** The test of the interaction between RSV and Flu A. **B.** The test of the interaction between RSV and Flu B.

## Discussion

In this study, we demonstrate the circulation dynamics of RSV and seasonal influenza in the U.S and examine their potential competition and mechanism at a population level. Our phylogenetic analyses initially suggest there might be a negative association between RSV and seasonal influenza by studying the correlation of their estimated time-series genetic diversity. We further study this potential negative interaction explicitly by applying a two-pathogen transmission model with HHS regional-level surveillance reports. Taken together, our analyses provide statistical support for the negative interaction between RSV and seasonal influenza after infection.

By studying the co-circulation of RSV and seasonal influenza in different regions of the U.S, we confirm the temporal correlation between these viruses. We found there might be little to zero number of weeks of time-lags to maximize the correlation coefficient between RSV and different subtypes of influenza. These differences in relative epidemic peaks of RSV and seasonal influenza across different HHS regions in the U.S suggests they might interact with different severity.

Phylogenetic analysis can be used to estimate the virus’s genetic diversity through time and therefore has the potential to reflect the effects of interactions between viruses. Previous phylogenetic studies of seasonal influenza demonstrated a periodicity in the relative genetic diversity in temperate zones where regional epidemics have coinciding epidemic peaks and seasonal patterns [27]. A similar seasonality of RSV was also reported in multiple Northern Hemisphere countries experiencing a temperate climate, with the annual epidemics starting in the autumn, peaking in winter, and ending in spring [28]–[30]. Therefore, we expect a similar seasonal periodicity in relative genetic diversity could be observed in the RSV phylogenetic analyses with sequences that are collected from available northern hemisphere countries. However, our phylogenetic analyses of RSV suggest a different pattern of seasonal influenza and indicate the potential of regional persistence, which is consistent with previous phylogenetic studies of RSV in other regions [31]-[32], [33]. The antigenic drift of seasonal influenza creates competition among strains, resulting in the emergence of selective forces and previously circulating strains can be prone to local extinction each year [34]. This negative selection including the effect of the vaccine causes rapid influenza population turnover and therefore it only takes a few years for contemporaneous strains to find a common ancestor [35], [36]. In contrast, RSV phylogenies harbor many deep branches in our analysis (Fig. 2). This reconstructed population dynamics of RSV might indicate latency supported by low levels of circulation during the summer season that is not captured by current RSV genomic surveillance [33], and RSV seasonal epidemics could be driven by other ecological factors like humidity, temperature, and human contact or travel [28][37]. These differences between RSV and influenza suggest in phylogenies suggest different protection and vaccine strategies may be needed against these viruses [34][35], [36]. We observe a negative correlation between RSV and seasonal influenza lineages with time series effective population sizes that are estimated from genetic data, which suggests a potentially negative interaction between these viruses on an ecological scale. However, our analyses are limited by the lack of genetic surveillance effort and more explicit analyses are still needed to study the potential competition between RSV and seasonal influenza.

Our transmission modeling work provides additional evidence of the negative interaction between RSV across regions and seasonal influenza and further demonstrates the potential mechanism of interaction. We find adding inhibition on co-infection to the model has relatively small effects on epidemic trajectories from a simulation study (Fig.3), and the inhibition on co-infection is not supported by the surveillance data from some HHS regions. This might be due to the short infection period of RSV and seasonal influenza, and only a small proportion of the population has the potential to be infected by two viruses simultaneously and therefore do not leave a strong dynamical footprint in population-level weekly surveillance data. In our analyses, there are very few individuals to be infected by RSV and seasonal influenza simultaneously, which is consistent with previous surveillance studies that suggest the observed incidence of co-infectivity of RSV and influenza was significantly less than expected [9]. To further confirm our estimation, the exact number of cases with co-infection is needed. In addition, the severities of competition between these viruses across HHS regions are estimated to be different from our analysis. The population structure (e.g. age) in different regions might cause different levels of cross-pathogen immunity [18]. It is also important to note that the surveillance reporting institutions vary across regions, which may result in different levels of competition that are reflected in the surveillance data. These location-specific features need to be considered when generalizing the findings.

Much of the previous evidence for the interaction between RSV and seasonal influenza is on an individual, biological level [38][25][24][39][21]. Previous evidence of the interaction of RSV and seasonal influenza on the population level implies that prevention of one could inadvertently lead to an increase in the burden of the other [23][26]. Recent modeling work suggests a competition between RSV and influenza with infection reducing heterologous acquisition by 41% for 10 days among children after infection in Nha Trang, Vietnam [40]. We provide additional evidence for the competition between RSV and influenza at the population level in a more diverse region in the U.S. and the level of competition could be differentiated by location. While our findings are consistent with previous experimental studies and modeling work suggests a competitive interaction between RSV and influenza [20][21][41][42], it is important to emphasize social behavioral changes after infection (sequestration during convalescence) with a virus may also play a role [43].

To simplify the analyses with the transmission model, we assume RSV-influenza interaction is bidirectional and the strength and duration of interaction that influenza exhibits on RSV are the same and vice versa. We do not model RSV and two subtypes of seasonal influenza together because adding another virus would significantly increase the complexity of the model and we believe there is a low chance for individuals to be infected with RSV and two subtypes of seasonal influenza within a short time. To reduce the number of parameters that need to be estimated, we fix the cross-protection period with a relatively long time of 6 months [44] and we assume the cross-immunity could protect the individual to be less likely infected by another pathogen at the same season. Our analysis is also limited by current RSV and seasonal influenza surveillance availability. We are lacking RSV subtype information in the current surveillance support. In addition, the reporting institutions of RSV and seasonal influenza surveillance vary between years, and spikes in detections may reflect increased testing within a given surveillance year. Hence, additional high-quality surveillance data in the future might be able to draw a more reliable conclusion.

Overall, we characterize the seasonal overlaps and evolutionary dynamics of RSV and seasonal influenza and focus on their potential interaction. Our work highlights the use of mathematical mechanistic models to test the interaction hypothesis of RSV and seasonal influenza. Although more effort is needed, our results suggest a cross-immunity-induced negative interaction of RSV and influenza on the ecological scale and can be identified in surveillance data. This study is helpful to have a better understanding of the different dynamics and the potential interaction between RSV and seasonal influenza, which are critical for predicting the effects of alteration of their ecological balance and designing vaccines against these viruses.

## Materials and Methods

### RSV and seasonal influenza surveillance data

PCR reports from HHS regional-level surveillance data for RSV that are collected from 2011 to 2019 were requested from CDC’s National Respiratory and Enteric Virus Surveillance System (NREVSS). HHS regional-level type-specific seasonal influenza (Flu A / Flu B) positive test reports were downloaded from FluView Interactive [45], and the viral surveillance reported by Clinical Labs was used after the year 2015. The national-level surveillance data were aggregated from the reports of 10 HHS regions.

### Correlation coefficient estimation

To measure the association between RSV and seasonal influenza epidemic, we computed correlation coefficients with weekly surveillance reports of different pairs of viruses in each HHS region in R v4.1. The weekly positive cases / million individuals of each virus were calculated by the estimated population for each HHS region in the given year, which were collected from the U.S. Census Bureau using the “State Population Totals and Components of Change: 2010-2019” data set [46]. We employed the Shapiro-Wilk normality test to test for the normality of the error distribution of the time series being compared. Pearson’s correlation coefficient was used to quantify the linear association among the co-circulating viral dynamics and the association was taken to be statistically significant at a p-value < 0.05 in a t-test. The time-lagged correlation coefficient was estimated in R package astsa v1.15 [47].

To examine the correlation of the genetic diversity for RSV and seasonal influenza, approx() interpolating function in R was used to generate the effective population sizes of different virus strains at the same time points from 2011-2019 from their corresponding Bayesian “Skyride” analysis. After examining the normality of the data using Shapiro-Wilk normality test, we used Pearson’s correlation coefficient test to measure the correlation of RSV and seasonal influenza

### Genetic datasets

HA segments of seasonal influenza (H3, H1, Victoria, and Yamagata) were retrieved from the GISAID database spanning the 2011-2019 epidemic seasons (generally considered as the October of the start year through May of the following year) in the U.S. Due to the lack of sampling for RSV in the US and North America regions, RSV G gene sequences (at least 70% length of complete G gene) that are collected from available northern hemisphere countries during the same period were retrieved from the GISAID database. RSV genotypes were determined with a reference dataset using NCBI BLAST [48], [49] and further confirmed with phylogenetic analyses, and sequences belonging to the dominant genotypes “ON” (RSV-A) and “BA” (RSV-B) were selected for the following analysis.

Nucleotide sequences that belong to each genotype were aligned using MAFFT.v7 [50] separately and initial maximum likelihood (ML) phylogenetic trees were built using RAxML.v8 [51]. The temporal signal of each dataset was analyzed using TempEst v1.5.3 [52] and temporal outliers were removed for further studies. These ML phylogenies for seasonal influenza in different subtypes were further used to subsample to a maximum of 100 isolates in each season using the Phylogenetic Diversity Analyzer (PDA) [53]. These subsampled datasets were used for the subsequent Bayesian phylogenetic analyses. The final datasets contained 1004 ON1, 769 BA RSV G gene sequences, 867 H3, 871 H1,796 Victoria, and 789 Yamagata HA gene segments of seasonal influenza.

### Bayesian phylogenetic analysis

Evolutionary dynamics of RSV and seasonal influenza were estimated with a Bayesian phylogenetic approach using BEAST v.1.10.4[54]. A generalized time-reversible nucleotide substitution model with gamma rate heterogeneity was used for the analysis of RSV and the SRD06 codon position model was used for the analysis of seasonal influenza [55][56].

Uncorrelated lognormal relaxed clock and GMRF Skyride coalescent model [57] were used to estimate the population dynamics of the virus during each season for both pathogens. At least 4 independent MCMC chains of 150-200 million generations were simulated to ensure a sufficient effective sample size (ESS > 200) as diagnosed in Tracer v1.7 [58]. LogCombiner v1.10.4 was used to combine the multiple runs and the Maximum Clade Credibility (MCC) tree was summarized in TreeAnnotator v1.10.4 after the removal of the 10% burn-in. Temporal-sliced descriptive statistics, including TMRCA estimates, diversity measurements, and mean statistics including coalescent rate, diversity, and Tajima’s D scores were calculated from BEAST sampled Bayesian phylogenetic trees using the software package PACT v0.9.3 [35].

### Two pathogen transmission model

We applied a two-pathogen transmission model to study the potential interaction of RSV and seasonal influenza [59]. The population is compartmentalized into Susceptible (S), Infectious (I), Cross-protected (C), and Recovered (R) for each pathogen 𝑖, (𝑖 𝜖 1,2)[59], [60]. Transitions among these compartments were governed by the following processes. Susceptible individuals with no immunity are infected at rates 𝜆_𝑖_ and enter the corresponding I compartment. Following the infectious period (with a mean duration of 1/ 𝛾_𝑖_), individuals recover and move to the C compartment, where individuals are fully protected against homologous re-infection and have some cross-immunity against another virus. Individuals then lose this cross-immunity but retain pathogen-specific immunity in moving to the R compartment at rate 𝜌. Pathogen-specific immunity wanes as individuals become susceptible again at rate 𝜔. The birth-death rate at each compartment is 𝜇.

We hypothesize that the infection with another pathogen might be less likely either during or after the infection of the first pathogen, corresponding to either the I or C compartments, and therefore we define the infection rate of another pathogen with the potential of reduction. In the I compartment, an interaction corresponds to a reduction in the infection rate of the second pathogen at 𝜓𝜆_𝑖_, where the inhibition to be co-infected is modeled by scaling the rates of infection using a value between 0 and 1 (𝜓 𝜖 [0,1]). In the C compartment, we similarly model a reduced infection rate 𝜒𝜆_𝑖_ for the second virus, wherein the strength of cross-immunity, 𝜒, takes a value between 0 and 1 (𝜒𝜖 [0,1]). Parameters and estimated values used in the model are shown in SI Appendix, Table S4. In a deterministic setting, the model is described by 16 ordinary differential equations (Table S3, Equations for each state can be read directly from SI Appendix, Fig. S3).

The inhibition of co-infection was modeled by a reduction in the infection rate to the second virus, 𝜓𝜆_𝑖_ (𝜓 𝜖 [0,1]), the equations governing these dynamics are given by:

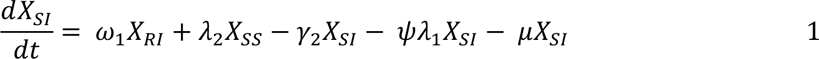

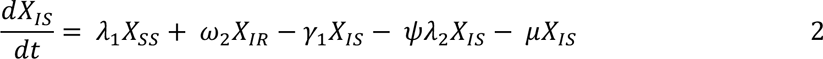

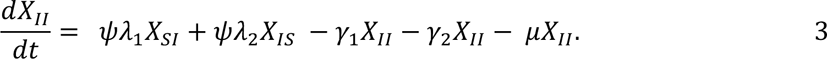

The cross-immunity after the infection was modeled by a reduced infection rate to the second virus 𝜒𝜆_𝑖_ (𝜒 𝜖 [0,1]) at the C compartment:

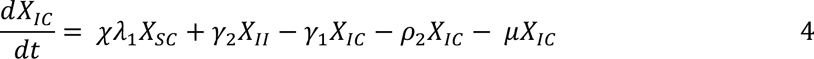

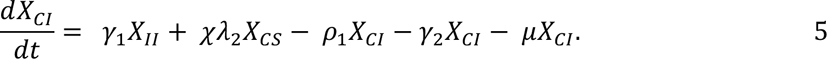

The overall population in this model is the sum of the populations in each compartment:

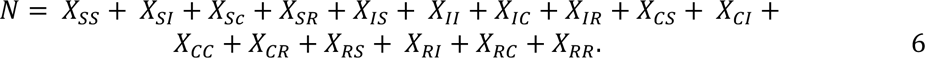

For virus 𝑖 (𝑖 ∈ {1,2}) in this model, seasonality in the transmission rate (𝛽_𝑖_(𝑡)) was incorporated in the transmission model, where 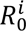 is the basic reproductive number, 𝑏_𝑖_ is the amplitude of seasonality, and 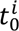 is the peak transmission day during the season:

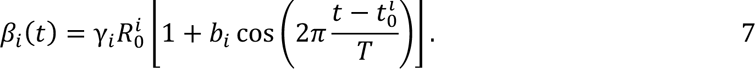

The force of infection depends on the number of individuals infected with the virus 𝑖 and the rate at which cases are imported into the population from external sources 𝜂_𝑖_.

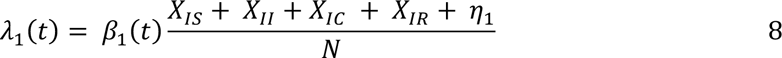

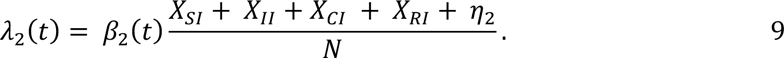

The deterministic model was implemented in R using the package “pomp” [61]. For the likelihood-based inference, we generated the observed cumulative number of cases (𝛫_𝑖_) according to the following equations, where 𝛿_𝑖_ is the case report rate of the virus.

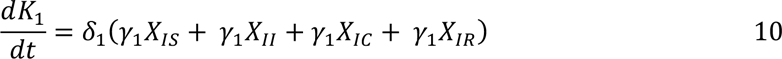

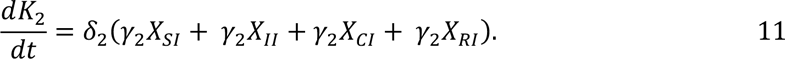

We initiated the model with an endemic equilibrium of the non-interacting model (𝜓 = 1, 𝜒 = 1) without seasonality (𝑏_1_ = 0, 𝑏_2_ = 0), the first 100 years were discarded as transient dynamics before the likelihood calculation.

### Hypothesis testing and parameter estimation

The model was fit with RSV and type-specific seasonal incidence surveillance that are collected in 10 HHS regions from 2014 to 2017. Maximum-likelihood estimates (MLEs) for the unknown parameters were found using trajectory matching. Differential evolution algorithm (DEoptim) for global optimization implemented in R package “DEoptim” [62] was used to find the estimates that maximize the MLE. We calculated AIC using MLE estimates under each hypothesis. If a model is more than 2 AIC units lower than another, we then consider it significantly better. RMSE was calculated to quantify how well the model fits with surveillance data. As proof of concept, we performed an out-of-sample simulation to demonstrate the suitability of the MLE model; that is, we generated the forecast for the following season of the MLE model with the estimated parameters and then compare the simulated cases with RSV and seasonal influenza surveillance that are collected in each HHS region.

We performed a simulation study to estimate the uncertainty of the competitive interaction parameters. We created likelihood profiles for the respective parameters by fitting a smooth line through the log of repeated likelihood estimates in which the respective parameter is a fixed value. The 95% CI is taken to 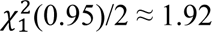 log-likelihood units below the maximum likelihood estimates using the 𝜒^’^distribution.

## Data availability

Source code and data have been deposited on GitHub: https://github.com/JianiC/RSV_flu The GISAID accession number of RSV and seasonal influenza sequence of this study are available in supplementary materials.

## Acknowledgments

We acknowledge the RSV surveillance data from the NREVSS team and subtype-specific seasonal influenza surveillance data from the CDC’s FluView Interactive. We acknowledge the originating and submitting laboratories for our use of sequences from the GISAID’s EpiFlu and EpiRSV Database.

